# Population behavioural dynamics can mediate the persistence of emerging infectious diseases

**DOI:** 10.1101/2021.05.03.21256551

**Authors:** Kathyrn R Fair, Vadim A Karatayev, Madhur Anand, Chris T Bauch

## Abstract

The critical community size (CCS) is the minimum closed population size in which a pathogen can persist indefinitely. Below this threshold, stochastic extinction eventually causes pathogen extinction. Here we use a simulation model to explore behaviour-mediated persistence: a novel mechanism by which the population response to the pathogen determines the CCS. We model severe coronavirus 2 (SARS-CoV-2) transmission and non-pharmaceutical interventions (NPIs) in a population where both individuals and government authorities restrict transmission more strongly when SARS-CoV-2 case numbers are higher. This results in a coupled human-environment feedback between disease dynamics and population behaviour. In a parameter regime corresponding to a moderate population response, this feedback allows SARS-CoV-2 to avoid extinction in the trough of pandemic waves. The result is a very low CCS that allows long term pathogen persistence. Hence, an incomplete pandemic response represents a “sour spot” that not only ensures relatively high case incidence and unnecessarily long lockdown, but also promotes long-term persistence of the pathogen by reducing the CCS. Given the worldwide prevalence of small, isolated populations in which a pathogen with low CCS can persist, these results emphasize the need for a global approach to coronavirus disease 2019 (COVID-19) vaccination.

## 1 Introduction

In classical mathematical models of acute infections that generate immunity, the introduction of a novel pathogen with a basic reproduction number *R*_0_ *>* 1 in a susceptible population typically causes a sharp initial spike whereby a very large proportion of the population is infected due to the lack of pre-existing immunity [1]. This is followed by a deep and prolonged trough during which there is a high risk of pathogen extinction due to stochastic effects when the number of infected persons becomes small [2, 3].

However, many mechanisms can save a pathogen from stochastic extinction, and this is one reason why many novel emerging pathogens have become endemic in human populations. For instance, spatial heterogeneity can ensure that the epidemic persists globally as long as the epidemics are not synchronized between the patches, such that the pathogen activity is quiescent in some patches while being active in others [4, 5]. Similarly, host refuges–such as an animal reservoir where the pathogen can persist even during periods of extinction in human populations–can promote persistence [6, 7], as can pathogen evolution [7].

In the deterministic approximation, pathogens that survive this initial extinction risk enter a regime where epidemic dynamics are less extreme. The pathogen may still become extinct in sufficiently small populations over a longer time period due to stochastic fade-out. In a sufficiently large population, however, chains of transmission can be sustained almost indefinitely, leading to long-term persistence of the pathogen [8]. The critical community size (CCS) is the minimum closed population size in which a pathogen persists indefinitely. Below this threshold, stochastic fade-out may occur [9–12]. An infection with a low CCS is more difficult to eradicate globally, since it can persist in small pockets of population. Hence, estimation of CCS is useful for determining the feasibility of pathogen control, elimination, and eradication strategies.

Mathematical models are often leveraged to explore CCS, since the likelihood of populations of varying sizes experiencing stochastic fade-out can be determined by running simulations [11, 13, 14]. Here we suggest a new mechanism by which a novel pathogen can avoid stochastic fade-out. Human populations respond to the presence of a novel emerging pathogen by adopting mitigation efforts. However, populations are complex adaptive systems [15] and thus may not respond vigorously enough to eliminate the infection right away [16]. And yet, for a sufficiently virulent pathogen, the population response may be strong enough to dampen transmission [17]. Since immunizing vaccines and other pharmaceutical interventions are rarely available when a novel pathogen emerges, populations rely upon scalable non-pharmaceutical interventions (NPIs) like physical distancing, mask use and hand washing. Hence, by preventing herd immunity in the population through infection, pathogen persistence is facilitated by stimulation of a behavioral response that ensures a sustainable reservoir of susceptible individuals. At the same time, if the population response relaxes when cases of infection fall to low levels, infection rates can rebound and stochastic fade-out can be prevented. We hypothesize that this mechanism of ‘behaviour-mediated persistence’ can support persistence of the pathogen by significantly reducing the critical community size.

This mechanism may be relevant to severe coronavirus 2 (SARS-CoV-2) transmission, where scalable NPIs have been highly successful in preventing cases and deaths [17, 18]. Here we develop a stochastic model of SARS-CoV-2 transmission in a population. We use it to illustrate how behaviour-mediated persistence can strongly reduce the CCS for SARS-CoV-2. The model includes both individual and institutional behavioural feedback on disease dynamics through individual adherence to NPIs and school/workplace closure. Such coupled human-environment models have been widely explored in the literature [19–21], including for COVID-19 [22–27]. Our objective is to illustrate the robust effect of behaviour-mediated persistence in the context of an epidemic caused by a novel emerging pathogen that causes acute immunizing infections in humans. We show that behavioural feedbacks of similar intensity to those observed during the COVID-19 pandemic can result in very small critical community sizes, thus promoting long-term persistence of the virus in human populations.

## 2 Model

Our stochastic compartmental model of SARS-CoV-2 transmission incorporates testing, super-spreaders, individual adherence to NPIs, and closing schools and workplaces. Disease progressions follows an *SEPAIR* pathway: susceptible to infection (*S*), exposed i.e. infected but not yet infectious (*E*), pre-symptomatic and infectious (*P*), asymptomatic i.e. infectious without developing symptoms (*A*), infectious i.e. symptomatic and infectious (I), or removed i.e. no longer infectious (*R*). Testing of symptomatic individuals occurs, and has an associated daily probability of the result becoming known. These individuals have either a known (*K*) or unknown (*U*) infection status.

An individual’s state, {*D*_*i*_, *T*_*i*_}, is based on their epidemiological, *D*_*i*_ *∈* {*S, E, P, A, I, R*}, and testing, *T*_*i*_ *∈*{*U, K*}, status. Individuals with epidemiological status *D*_*i*_ and either testing status are included within {*D*_*i*_, ·}, likewise for {·, *T*_*i*_}. On day *t*, the number of individuals with state {*D*_*i*_, *T*_*i*_} is 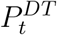, and *P*_*t*_ is the total population size. Every day begins with births (addition of susceptible individuals) and non-COVID-19 deaths (removal of individuals from all states). Following this, individual’s states are updated to mark their progression through infection states and to indicate their testing status (See Supplementary methods for details).

Infection probabilities are impacted by: (1) an individual’s contacts in schools, workplaces, homes, and other locations; (2) the extent to which closures and individual adherence to NPIs reduce transmission; (3) the effect of population size on transmission rates; and (4) super-spreading (as detailed in Supplementary methods). We set the fraction of an individuals contacts occurring in schools and workplaces to be *w* = 0.45; these contacts are reduced through closure of these locations [28]. The remaining fraction of contacts (1 − w) occur in homes and other locations where individual adherence to NPIs reduces contacts. Reduction of contacts through individual NPI adherence occurs in proportion to the prevalence of confirmed positive cases within the population 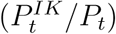 The fraction of contacts, *F* (*t*), remaining on day *t* after accounting for the impacts of closures and individual NPI adherence is

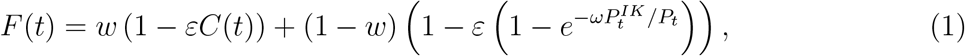

where *ε* is the efficacy of NPIs (closures, individual-level measures such as mask-wearing, etc.) in reducing contacts. Returning a positive COVID-19 test will cause an individual to reduce their contacts by a fraction η, the effect of which is incorporated using *f*_*T* =*U*_ = 1, *f*_*T* =*K*_ = 1 − *η*, where *η* = 0.8 [29, 30]. A susceptible individual’s daily probability of becoming infected is

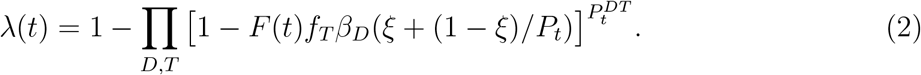

Here *ξ* controls how transmission probability is influenced by population size: *ξ* = 1 returns mass-action incidence and *ξ* = 0 returns standard incidence [31, 32].

The initial 90-day closure (of schools and workplaces) begins when *t* = *t*_*close*_; the first day that the cumulative number of confirmed cases divided by *P*_*t*_ exceeds γ (the threshold prevalence triggering a closure). All subsequent closures are enacted based on the current prevalence of confirmed cases, with the condition 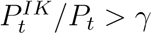. These closures last for δ_*C*_ = 30 days, with *t*_*C*_ indicating the day the last closure was enacted. Decisions are re-evaluated after the expiration of a closure period. We define the closure function, *C*(*t*), for this strategy as

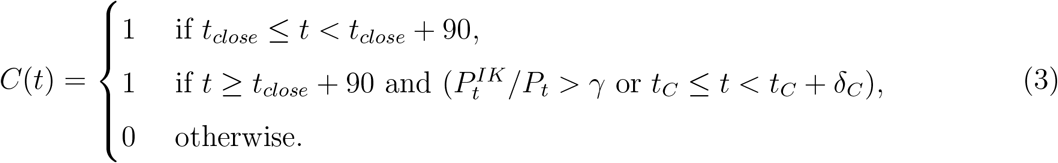

Table S1 contains all parameter definitions, baseline values, and sources. Simulations, analysis, and visualization are performed using R (Version 4.0.3) in Rstudio (Version 1.2.5019) [33,34]. All code is available in a GitHub repository [35]. See Supplementary methods for details of how simulations are implemented, and calculations used to determine the CCS and the proportion of the population infected.

## 3 Results

### Baseline scenario

Our baseline scenario is a situation of moderate NPI control: NPIs maintain SARS-CoV-2 case incidence at a relatively low level, and in some stochastic realizations they are even able to eliminate it, in a closed population of 50,000 individuals over a 1-year period. This is achieved through an initial closure of schools and workplaces (‘lockdown’), accompanied by additional episodes of closure that are triggered when infection prevalence thresholds are exceeded (Fig. 1). For comparison, a scenario of strong NPI control appears in the Supplementary Appendix (Fig. S1), along with three additional scenarios showing the effect of having no response, or a weak response either on the part of the public health authorities, or the population (Fig. S2-S4). Starting from this baseline, we explore how changes in parameter values for human behaviour, testing, and transmission can inhibit or facilitate the pathogen persistence, or shift outcomes to improve or decrease the effectiveness of NPIs.

**Figure 1.**
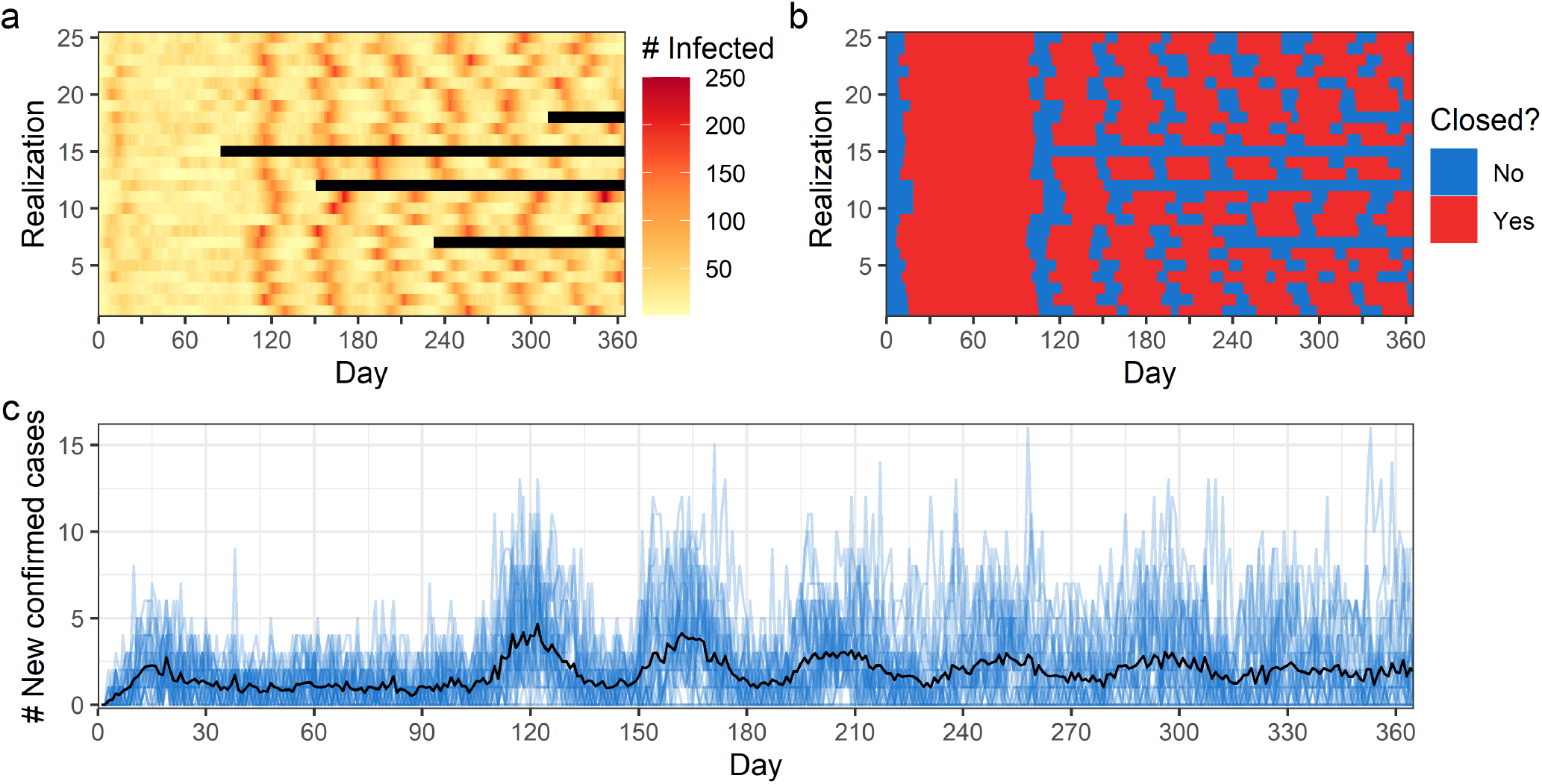
Baseline scenario: moderate NPI response by government and population. Figure panels show (a) number of infected individuals within the population (black indicates zero infected) (b) whether closures are in place and daily number of (c) new confirmed cases. In (a) and (b) each row corresponds to a single realization, while in (c) blue lines indicate each realization, with the black line indicating the mean outcome across all realizations. Model parameter settings are shown in Table S1.

### Moderate control efforts reduce the critical community size

A scenario where neither the population nor the government does anything causes high infection rates, followed by burn out of the pathogen after most of the population becomes infected and immune (or deceased) (Supplementary Fig. S2). On the other hand, a strong NPI response can result in small numbers of infections, and thus the pathogen is eliminated through stochastic fade out (Supplementary Fig. S1). However, a moderate NPI response avoids of these extremes through a ‘slow burn’ where moderate rates of infection are sustained over long periods of time, thereby enabling persistence (Fig. 1). Across a range of parameter values, we observe that the CCS for intermediate efficacy of school and workplace closure (*ε*) and intermediate risk perception among members of the population (*ω*, a driver of individual NPI adherence), is actually smaller than when *ω* and *ε* are very high or very low (Fig. 2, Fig. 3).

**Figure 2.**
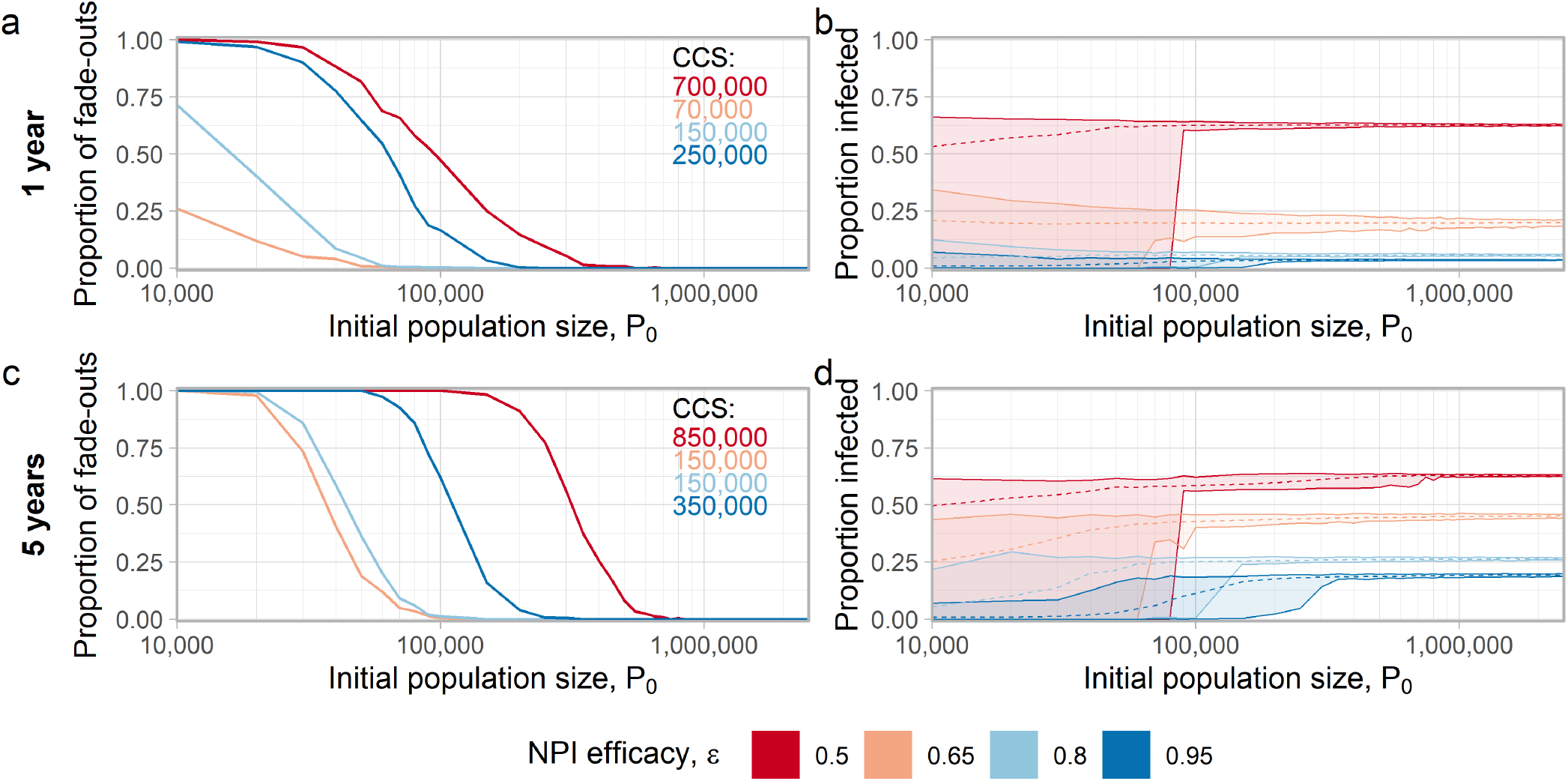
Critical community size is smallest for intermediate NPI efficacy. Very high or very low NPI efficacy (*ϵ*) generates a large CCS. Very low *ϵ* generates the largest number of infections. Figure panels show (a) the proportion of fade-outs and (b) the mean proportion of the population infected (dashed lines) after 1 year. Results after 5 years are shown in (c) and (d). In (a) and (c) inset panels indicate critical community sizes, and in (b) and (d) ribbons display minimal and maximal values across all realizations. Settings for all parameters (except *ε*) are shown in Table S1.

**Figure 3.**
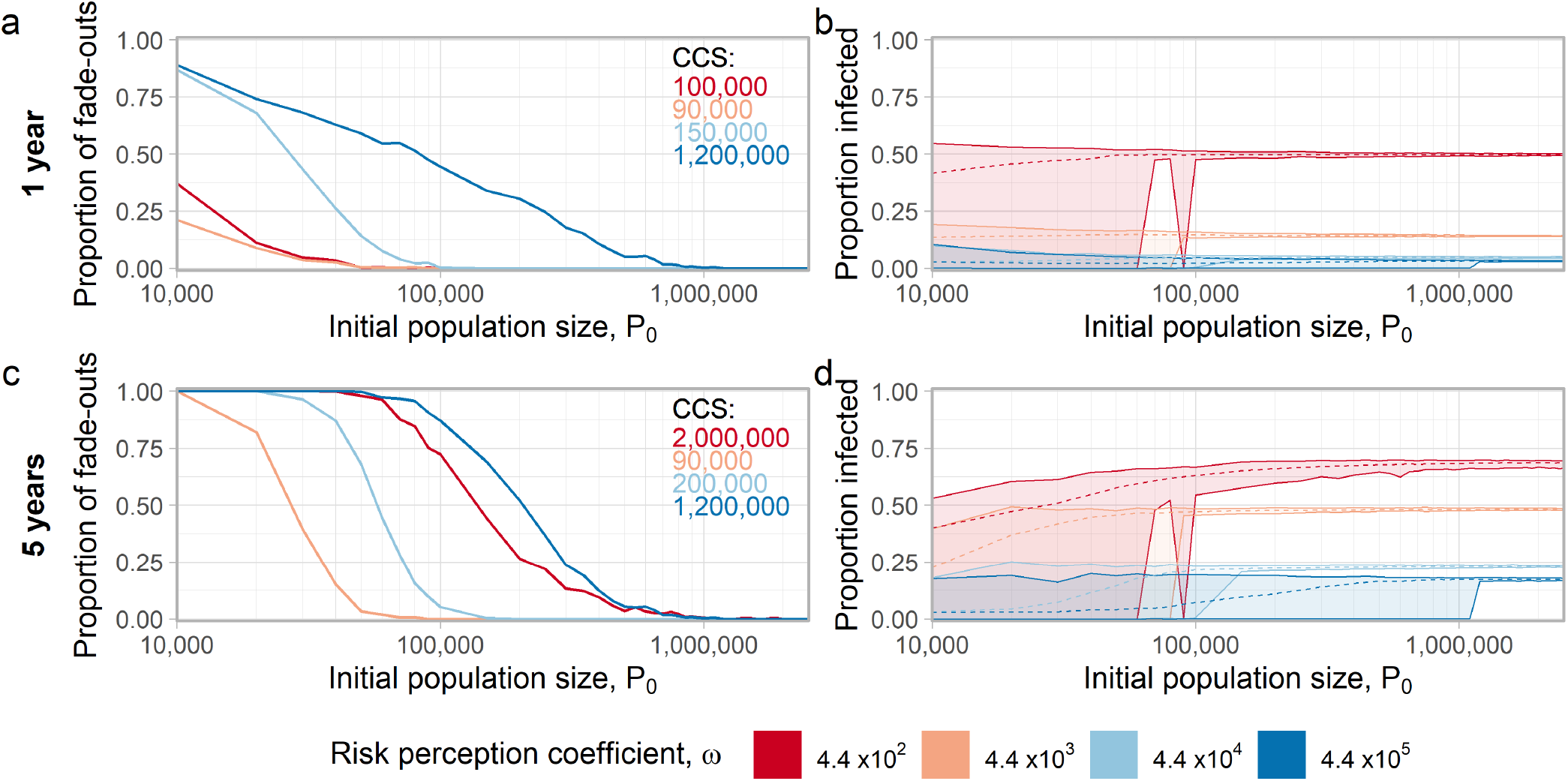
Critical community size is smallest for intermediate population risk perception. Very high or very low risk perception (*ω*) results in a large CCS. Very low *ω* generates the largest number of infections. Figure panels show (a) the proportion of fade-outs and (b) the mean proportion of the population infected (dashed lines) after 1 year. Results after 5 years are shown in (c) and (d). In (a) and (c) inset panels indicate critical community sizes, and in (b) and (d) ribbons display minimal and maximal values across all realizations. Settings for all parameters (except *ω*) are shown in Table S1.

Compared to small *ε*, moderate values of *ε* will result in a smaller proportion of individuals becoming infected, but also reduce the critical community size by several hundred thousand individuals (Fig. 2). Moderate *ε* causes small populations to experience persistent outbreaks over a longer period of time than they would if there was either a weak or a strong response. We emphasize that the larger CCS obtained for a low *ε* comes at the cost of a higher number of infections (Fig. 2b,d). Only a high value of *ε* can achieve both a large CCS and small number of cases. The proportion of fade-outs shows a similar overall pattern, with both low and high values of *ε* associated with a high rate of stochastic fade-out, on account of burnout and effective control, respectively (Fig. 2a,c).

The parameter *ω* (which governs the effort that individuals devote to NPI adherence in response to their perceived individual risk of contracting SARS-CoV-2) has a very large impact on the critical community size (Fig. 3). The role of moderate risk perception in reducing the CCS is apparent over a 5-year time period but not a 1-year period. Over a 1-year time period, the CCS is relatively low for both low and moderate *ω* (90,000 to 150,000). Also, the proportion infected is very high at the lowest value of *ω*. However, when *ω* is very high and thus individuals engage in strict adherence to NPIs even when confirmed case prevalence is low, the CCS is 1,200,000.

For a 5-year horizon, both very high and very low risk perception values increase the CCS by at least 1,000,000 individuals as compared to a case of moderate risk perception. However, when risk perception is low this comes at the cost of more than half of the population becoming infected. As with other parameters related to human behaviour, there are clear diminishing returns on increases in risk perception for a 1-year horizon. Outcomes over a 5-year horizon depend on the initial population size *P*_0_, as expected.

### Impact of transmission assumptions

The parameter *ξ* controls whether the risk of a person becoming infected depends upon the total number of infected individuals (mass action incidence, *ξ* = 1) or whether it depends on the population density of infected individuals (standard incidence, *ξ* = 0). This parameter impacts both the critical community size and the cumulative number of infections. When *ξ* = 0, the cumulative number of infections relatively low, as is the CCS (Fig. 4). When *ξ* is small (*ξ* = 1 ×10^−5^) a clearly defined CCS does not even exist, because fade-outs occur not only when population sizes are small, but also when they are sufficiently large, due to burnout.

**Figure 4.**
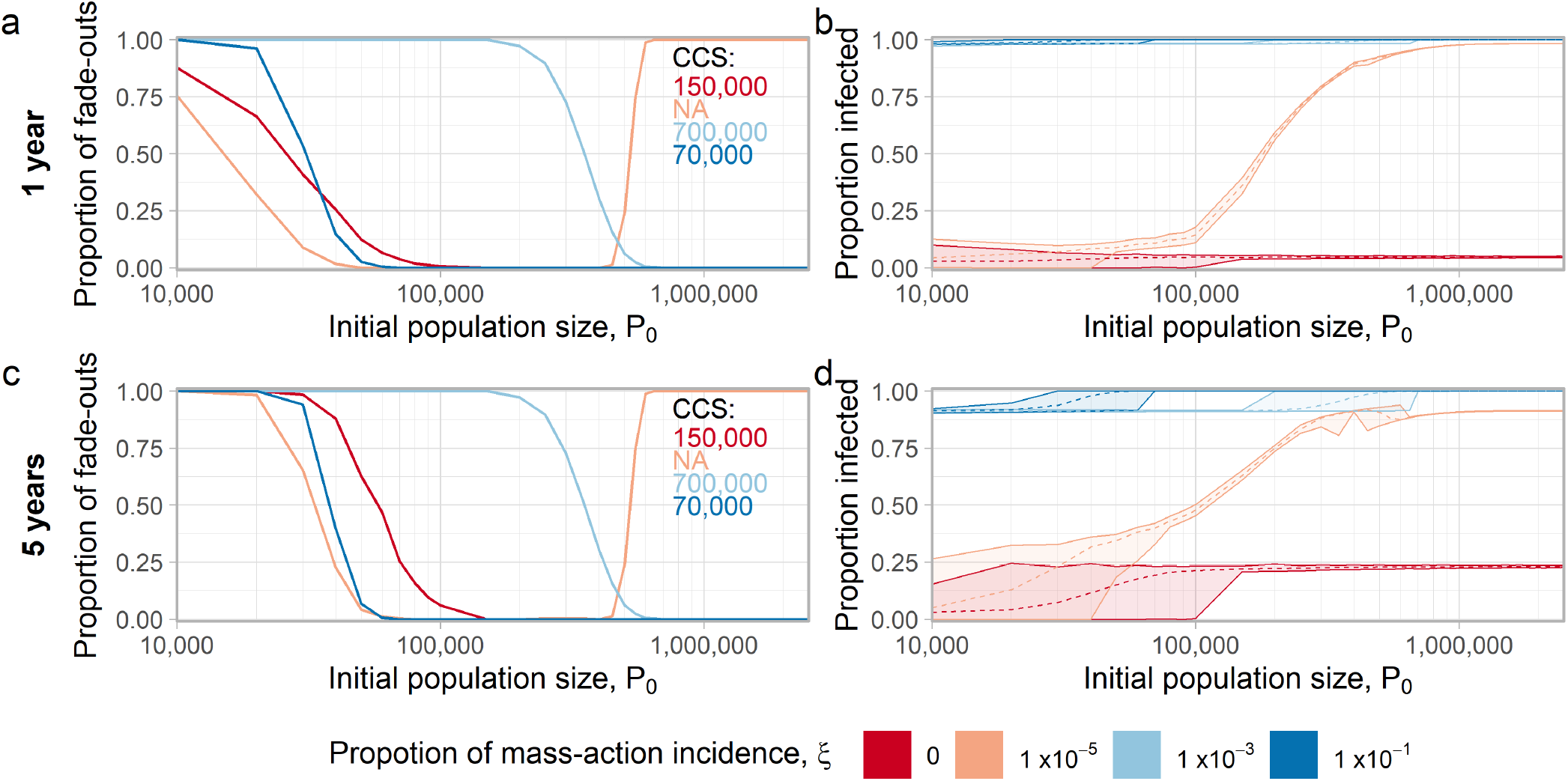
Both the CCS and number of infections are strongly influenced by mass-action versus standard incidence. Figure panels show (a) the proportion of fade-outs and (b) the mean proportion of the population infected (dashed lines) after 1 year. Results after 5 years are shown in (c) and (d). In (a) and (c) inset panels indicate critical community sizes, and in (b) and (d) ribbons display minimal and maximal values across all realizations. Settings for all parameters (except *ξ*) are shown in Table S1.

When the level of mass action incidence is moderately high or high (*ξ* ≥ 1 ×10^−3^), the virus infects the majority of the population and the mixture of mass-action and standard incidence results in a large CCS, whereas at *ξ* = 1 ×10^−1^ transmission occurs so rapidly that the virus persists in all but the smallest population sizes, leading to a small CCS.

As expected, an increased transmission probability reduces the CCS in our baseline scenario of moderate population NPI response, across a broad range of population sizes (Fig. S5). If the population size is above approximately 400,000 individuals, then stochastic fade-out does not occur over a 5-year period, even for the lowest transmission probability. For an intermediate population size range from 30,000 to 400,000, however, the chance of stochastic fade-out depends strongly on the transmission probability. We note that in the absence of a population NPI response, certain values of the transmission rate could be high enough to rapidly confer herd immunity to the population, and thus cause pathogen burn out.

Over a 1-year horizon, birth and death rates (Fig. S6, S7) do not substantially impact CCS values or the proportion of the population that becomes infected. However, over a 5-year horizon the number of infections decreases slightly when the birth or death rate is very high. In the presence of baseline (moderate) interventions, accounting for births and deaths results in the pathogen persisting within the population longer than it would in a population of fixed size (Fig. S8).

### Impact of testing and lockdown thresholds

Interventions such as school/workplace closure and testing impact both CCS and the cumulative number of infections. Increasing the testing rate from τ_*I*_ = 0.1 to τ_*I*_ = 0.7 increases the CCS by making it easier to eliminate the virus (Fig. S9). However, increasing the testing rate exhibits diminishing returns in other respects: an increase in τ_*I*_ generates large reductions in the number of infections when τ_*I*_ is small but not when τ_*I*_ is already large, after both one and five years.

The infection prevalence γ that triggers lockdown has relatively less impact on the CCS (Fig. S10), except when the trigger prevalence is very high (γ = 1.5 ×10^−2^), in which case the CCS is much larger than at lower γ-values, on account of burnout that occurs with a lenient shutdown threshold. As expected, higher values for trigger prevalence result in a larger cumulative number of infections. At a 1-year horizon, the duration of closures has little impact on the CCS and the proportion of the population infected (Fig. S11), though longer closures increase the probability of a fade-out occurring for *P*_0_ <150, 000. At a 5-year horizon, longer closures reduce the number of infections, and employing closures of at least 60 days increases the CCS.

## 4 Discussion

Non-pharmaceutical interventions (e.g. closures, physical distancing, and mask use) are crucial to pandemic response, especially in the early stages before vaccines or treatments have been developed [36]. However, NPIs require broad population adherence in order to be successful. Our results illustrate a mechanism of “behaviour-mediated persistence” whereby imperfect population NPI adherence means a novel emerging pathogen can persist in smaller populations than would otherwise be the case, if the population uptake of NPIs were stronger or if the population did not respond at all. Behaviour mediated persistence makes elimination of the pathogen more difficult.

Taken together, these results suggest a modelled CCS for COVID-19 ranging from approximately 70,000 to 2,000,000 depending on parameter settings for the proportion of mass action incidence (Fig. 4), efficacy of NPIs (Fig. 2), and risk perception (Fig. 3). A high CCS (which makes it easier to eliminate the infection) is achieved for high testing rates, high perceived risk of infection; high NPI efficacy or low transmission rate, as expected. These situations also achieve a low cumulative number of infections and correspond to stringent infection control. A high CCS also occurs when risk perception or NPI efficacy are sufficiently low (Fig. 2, Fig. 3), but this occurs through rapid burnout and comes at the cost of a high proportion of infected individuals. Moderate levels of risk perception or NPI efficacy result in low CCS by enabling a regime of constant, moderate, sustained infection in the population that lies between the extremes of burnout and elimination (Fig. 2, Fig. 3).

Because smaller populations are more numerous (and often less connected through travel) than large urban centres, the low CCS due to behaviour-mediated persistence means that infections could persist in smaller and more remote populations around the world for a long period of time. In many lower-income countries, these are the same populations that also have limited access to pharmaceutical interventions, which underscores the need for using transmission-interrupting vaccines to achieve elimination or eradication, unless COVID-19 evolves to a state of low virulence [37].

We found that imperfect population NPI uptake can result in a slow burn where the virus neither depletes the susceptible population rapidly, nor is eliminated by a quick and robust response. This finding dovetails with the findings of other models including feedback of infection dynamics on behaviour, whereby individuals respond more strongly when infections are more prevalent [19, 38, 39]. However, we emphasize that these findings do not recommend a “herd immunity” strategy of doing nothing, because the number of cases is enormous in the model scenarios where there is no population response. Rather, the optimal scenario is a strict response that both results in a high CCS and few infections, as occurs in our model under conditions of a high testing rate, high perceived risk, and/or high efficacy of NPIs. Our results emphasize the need to act quickly and decisively, both for government-mandated measures (e.g. the prevalence that triggers a closure) and individual efforts (e.g. adherence to NPIs as driven by risk perception). A sufficiently quick and strong response can limit the scope for pandemic fatigue by allowing an earlier re-opening, and may limit economic damage of the pandemic [40]. This corresponds well with empirical analyses concluding that combined interventions are necessary to “flatten the curve” [41].

This mechanism is also relevant to virulence evolution. Theories of virulence evolution often focus on the complex ways in which the response of host biology determines the evolutionary fitness of strains with differing levels of virulence [42–44]. In our model, the impact of virulence on decision-making is expressed implicitly through the parameter *ω*, which determines how strongly the population responds to a given prevalence of infection [45]. More virulent strains can be expected to be associated with a higher value of *ω*, which in turn changes transmission rates, infection incidence, and the CCS–all features that can impact evolutionary fitness. For instance, low or intermediate virulence might be selected for if it puts the pathogen in the *ω* regime of lower CCS, which promotes long-term persistence. We suggest that the social response of the host is as important as the biological response, when NPIs are being applied to pandemic mitigation. In a related vein, we note that a moderate NPI response permits higher case counts than a strong NPI response, and thus provides more opportunities for a more virulent or more transmissible strain to emerge.

We studied a single population, but in real populations, the infection could be re-seeded by case imports from other populations. This has implications for long-term pathogen persistence [12]. We also assumed that immunity is lifelong, although immunity may wane over time [37]. And, the virus may be capable of immune escape [46]. These simplifying assumptions could impact model dynamics, and should be explored in future research.

These findings illustrate the social-epidemiological aspect of pandemics. Human behaviour can be a crucial factor in determining the critical community size for any pathogen that stimulates a prevalence-dependent population response, as occurs during a pandemic for instance.

## Data Availability

Code used for simulations and visualization, as well as data sets generated from simulations (required to reproduce visualizations) are available in a GitHub repository (https://github.com/k3fair/COVID-19-SinglePatch-model).

## Acknowledgements

This research was made possible by the facilities of the Shared Hierarchical Academic Research Computing Network (SHARCNET: www.sharcnet.ca) and Compute/Calcul Canada.

## References

1. Herbert W Hethcote. The mathematics of infectious diseases. SIAM review, 42(4):599– 653, 2000.

2. Linda JS Allen and Glenn E Lahodny Jr. Extinction thresholds in deterministic and stochastic epidemic models. Journal of Biological Dynamics, 6(2):590–611, 2012.

3. Tom Britton, Thomas House, Alun L Lloyd, Denis Mollison, Steven Riley, and Pieter Trapman. Five challenges for stochastic epidemic models involving global transmission. Epidemics, 10:54–57, 2015.

4. David JD Earn, Pejman Rohani, and Bryan T Grenfell. Persistence, chaos and synchrony in ecology and epidemiology. Proceedings of the Royal Society of London. Series B: Biological Sciences, 265(1390):7–10, 1998.

5. C’secile Viboud, Ottar N Bjørnstad, David L Smith, Lone Simonsen, Mark A Miller, and Bryan T Grenfell. Synchrony, waves, and spatial hierarchies in the spread of influenza. science, 312(5772):447–451, 2006.

6. David E Stallknecht and Justin D Brown. Wild birds and the epidemiology of avian influenza. Journal of wildlife diseases, 43(3 Supplement):S15–S20, 2007.

7. David JD Earn, Jonathan Dushoff, and Simon A Levin. Ecology and evolution of the flu. Trends in ecology & evolution, 17(7):334–340, 2002.

8. Linda JS Allen and Amy M Burgin. Comparison of deterministic and stochastic sis and sir models in discrete time. Mathematical biosciences, 163(1):1–33, 2000.

9. Maurice S Bartlett. The critical community size for measles in the united states. Journal of the Royal Statistical Society: Series A (General), 123(1):37–44, 1960.

10. Maurice S Bartlett. Measles periodicity and community size. Journal of the Royal Statistical Society: Series A (General), 120(1):48–60, 1957.

11. Matthew J Keeling and Bryan T Grenfell. Disease extinction and community size: modeling the persistence of measles. Science, 275(5296):65–67, 1997.

12. C Jessica E Metcalf, Katie Hampson, Andrew J Tatem, Bryan T Grenfell, and Ottar N Bjørnstad. Persistence in epidemic metapopulations: quantifying the rescue effects for measles, mumps, rubella and whooping cough. PloS one, 8(9):e74696, 2013.

13. Helen J Wearing and Pejman Rohani. Estimating the duration of pertussis immunity using epidemiological signatures. PLoS Pathog, 5(10):e1000647, 2009.

14. Matthew J Ferrari, Rebecca F Grais, Nita Bharti, Andrew JK Conlan, Ottar N Bjørnstad, Lara J Wolfson, Philippe J Guerin, Ali Djibo, and Bryan T Grenfell. The dynamics of measles in sub-saharan africa. Nature, 451(7179):679–684, 2008.

15. Simon Levin, Tasos Xepapadeas, Anne-Sophie Cr’sepin, Jon Norberg, Aart De Zeeuw, Carl Folke, Terry Hughes, Kenneth Arrow, Scott Barrett, Gretchen Daily, et al. Social-ecological systems as complex adaptive systems: modeling and policy implications. Environment and Development Economics, 18(2):111–132, 2013.

16. Timothy C Reluga. Game theory of social distancing in response to an epidemic. PLoS Comput Biol, 6(5):e1000793, 2010.

17. Roy M Anderson, Hans Heesterbeek, Don Klinkenberg, and T D’seirdre Hollingsworth. How will country-based mitigation measures influence the course of the covid-19 epidemicã The Lancet, 395(10228):931–934, 2020.

18. Kathyrn Fair, Vadim Karatayev, Madhur Anand, and Chris Bauch. Estimating covid-19 cases and deaths prevented by non-pharmaceutical interventions in 2020-2021, and the impact of individual actions: a retrospective model-based analysis. medRxiv, 2021.

19. Zhen Wang, Michael A Andrews, Zhi-Xi Wu, Lin Wang, and Chris T Bauch. Coupled disease–behavior dynamics on complex networks: A review. Physics of life reviews, 15:1– 29, 2015.

20. Lee-Ann Barlow, Jacob Cecile, Chris T Bauch, and Madhur Anand. Modelling interactions between forest pest invasions and human decisions regarding firewood transport restrictions. PloS one, 9(4):e90511, 2014.

21. Kirsten A Henderson, Chris T Bauch, and Madhur Anand. Alternative stable states and the sustainability of forests, grasslands, and agriculture. Proceedings of the National Academy of Sciences, 113(51):14552–14559, 2016.

22. Vadim A Karatayev, Madhur Anand, and Chris T Bauch. Local lockdowns outperform global lockdown on the far side of the covid-19 epidemic curve. Proceedings of the National Academy of Sciences, 117(39):24575–24580, 2020.

23. Jinyu Wei, Li Wang, and Xin Yang. Game analysis on the evolution of covid-19 epidemic under the prevention and control measures of the government. Plos one, 15(10):e0240961, 2020.

24. Marco A Amaral, Marcelo M de Oliveira, and Marco A Javarone. An epidemiological model with voluntary quarantine strategies governed by evolutionary game dynamics. Chaos, Solitons & Fractals, 143:110616, 2021.

25. Sansao A. Pedro, Frank T. Ndjomatchoua, Peter Jentsch, Jean M. Tchuenche, Madhur Anand, and Chris T. Bauch. Conditions for a second wave of covid-19 due to interactions between disease dynamics and social processes. Frontiers in Physics, 8:428, 2020.

26. KM Ariful Kabir and Jun Tanimoto. Evolutionary game theory modelling to represent the behavioural dynamics of economic shutdowns and shield immunity in the covid-19 pandemic. Royal Society open science, 7(9):201095, 2020.

27. Peter C Jentsch, Madhur Anand, and Chris T Bauch. Prioritising covid-19 vaccination in changing social and epidemiological landscapes: a mathematical modelling study. The Lancet Infectious Diseases, 2021.

28. Bureau of Labor Statistics. American time use survey - 2018 results, 2018. Retrieved on June 16, 2020 from https://www.bls.gov/news.release/pdf/atus.pdf.

29. FA Soud, MM Cortese, AT Curns, PJ Edelson, RH Bitsko, HT Jordan, AS Huang, JM Villalon-Gomez, and GH Dayan. Isolation compliance among university students during a mumps outbreak, kansas 2006. Epidemiology & Infection, 137(1):30–37, 2009.

30. Joel Hellewell, Sam Abbott, Amy Gimma, Nikos I Bosse, Christopher I Jarvis, Timothy W Russell, James D Munday, Adam J Kucharski, W John Edmunds, Fiona Sun, et al. Feasibility of controlling covid-19 outbreaks by isolation of cases and contacts. The Lancet Global Health, 2020.

31. MCD Jong, O Diekmann, and H Heesterbeek. How does transmission of infection depend on population sizeã epidemic models. Publication of the Newton Institute, pages 84–94, 1995.

32. Janis Antonovics, Yoh Iwasa, and Michael P Hassell. A generalized model of parasitoid, venereal, and vector-based transmission processes. The American Naturalist, 145(5):661– 675, 1995.

33. RStudio Team. RStudio: Integrated Development Environment for R. RStudio, PBC., Boston, MA, 2020.

34. R Core Team. R: A Language and Environment for Statistical Computing. R Foundation for Statistical Computing, Vienna, Austria, 2020.

35. Kathyrn R Fair and Vadim A Karatayev. Covid-19-singlepatch-model, 2021. GitHub repository. https://github.com/k3fair/COVID-19-SinglePatch-model.

36. T Thanh Le, Zacharias Andreadakis, Arun Kumar, R Gomez Roman, Stig Tollefsen, Melanie Saville, and Stephen Mayhew. The covid-19 vaccine development landscape. Nat Rev Drug Discov, 19(5):305–306, 2020.

37. Jennie S Lavine, Ottar N Bjornstad, and Rustom Antia. Immunological characteristics govern the transition of covid-19 to endemicity. Science, 371(6530):741–745, 2021.

38. Chris T Bauch and David JD Earn. Vaccination and the theory of games. Proceedings of the National Academy of Sciences, 101(36):13391–13394, 2004.

39. Frederik Verelst, Lander Willem, and Philippe Beutels. Behavioural change models for infectious disease transmission: a systematic review (2010–2015). Journal of The Royal Society Interface, 13(125):20160820, 2016.

40. Asli Demirguc-Kunt, Michael Lokshin, and Ivan Torre. The sooner, the better: The early economic impact of non-pharmaceutical interventions during the covid-19 pandemic. World Bank Policy Research Working Paper, (9257), 2020.

41. You Li, Harry Campbell, Durga Kulkarni, Alice Harpur, Madhurima Nundy, Xin Wang, Harish Nair, Usher Network for Covid, et al. The temporal association of introducing and lifting non-pharmaceutical interventions with the time-varying reproduction number (r) of sars-cov-2: a modelling study across 131 countries. The Lancet Infectious Diseases, 21(2):193–202, 2021.

42. Samuel Alizon, Amy Hurford, Nicole Mideo, and Minus Van Baalen. Virulence evolution and the trade-off hypothesis: history, current state of affairs and the future. Journal of evolutionary biology, 22(2):245–259, 2009.

43. Troy Day and James G Burns. A consideration of patterns of virulence arising from host-parasite coevolution. Evolution, 57(3):671–676, 2003.

44. Troy Day and Stephen R Proulx. A general theory for the evolutionary dynamics of virulence. The American Naturalist, 163(4):E40–E63, 2004.

45. Joe Pharaon and Chris T Bauch. The influence of social behaviour on competition between virulent pathogen strains. Journal of theoretical biology, 455:47–53, 2018.

46. Kai Kupferschmidt. New mutations raise specter of ‘immune escape’. Science, 371(6527):329–330, 2021.

