## Supplementary information for "Population behavioural dynamics can mediate the persistence of emerging infectious diseases"

### Supplementary methods

Within the model, individuals states are updated based on the following events:

1. Exposure: $\{S, \cdot\}$ individuals are exposed to SARS-CoV-2 with probability $\lambda(t)$, shifting to $\{E, \cdot\}$.
2. Onset of infectious period: $\{E, \cdot\}$ individuals become pre-symptomatic and infectious with probability $(1-\pi)\alpha$, shifting to $\{P, \cdot\}$. Alternatively, they become asymptomatic and infectious (with probability $\pi\alpha$), shifting to $\{A, \cdot\}$.
3. Onset of symptoms: $\{P, \cdot\}$ individuals become symptomatic with probability $\sigma$, shifting to $\{I, \cdot\}$.
4. Testing: $\{I, U\}$ individuals are tested with probability $\tau_I$ shifting to $\{I, K\}$.
5. Removal: $\{I, \cdot\}$ and $\{A, \cdot\}$ individuals cease to be infectious with probability $\rho$, shifting to $\{R, \cdot\}$.

With probability $m = 0.0066$ [1], individuals transitioning from the symptomatic and infectious state ($\{I, \cdot\}$) to the removed (i.e. no longer infectious) state are assigned as COVID-19 deaths. Individuals who have died due to COVID-19 do not factor into subsequent birth and non-COVID-19 death calculations. At the onset of infectiousness, newly infected individuals have a probability $s = 0.2$ of being assigned as super-spreaders [2]. We differentiate between super-spreaders and non-super-spreaders using subscripts ($s$ and $ns$ respectively), such that we have $P_s$, $P_{ns}$, $A_s$, $A_{ns}$, $I_s$, $I_{ns}$. Super-spreaders have their probability of infecting others increased by a factor of $(1-s)/s$.

Based on research suggesting that 44% of SARS-CoV-2 shedding occurs before symptom onset [3] and our assumed period of infectiousness we set the transmission probabilities for asymptomatic and pre-symptomatic individuals to be half that of symptomatic individuals ($\beta_0^A = \beta_0^P = 0.5\beta_0^I$). Applying the additional assumption that $R_0 = 2.3$ [4], we determine that $(0.56)(2.3) = \beta_o^I/0.67$ and thus the symptomatic transmission probability is $\beta_0^I \approx 0.86$ [5]. Non-super-spreaders ($\{P_{ns}, \cdot\}, \{A_{ns}, \cdot\}, \{I_{ns}, \cdot\}$) have a transmission probability $\beta_{D_{ns}} = \beta_0^D$, while super-spreaders ($\{P_s, \cdot\}, \{A_s, \cdot\}, \{I_s, \cdot\}$) obtain $\beta_{D_s} = \beta_0^D(1-s)/s$.

When simulating scenarios to determine the baseline, we generate 25 realizations of the model; each begins with 50,000 people of which approx. 0.01% have been exposed to SARS-CoV-2. For simulations exploring the impact of varying parameters, we generate 500 realizations of the model for each $P_0$, with approx. 0.01% of the population exposed to SARS-CoV-2 at the initial time-step. As the initial exposure probability is low, some realizations will have no initial infections and are excluded from our analysis. For $P_0 < 100,000$ we increment $P_0$ by $10,000$, with increments of $50,000$ for $P_0 \in [100,000, 1,000,000)$ and of $100,000$ for $P_0 > 1,000,000$.

The CCS values we state are calculated by finding the smallest initial population size meeting the condition that, at that initial population size and at all larger initial population sizes, no fade-outs occur. These values are approximate, partially because we sample $P_0$ at increments (as outlined above) and thus it may be that the initial population size at which this condition is first met lies between the $P_0$ values we consider. As well, the stochastic nature of our model means that re-running the experiments may yield slightly different CCS values. Running the experiments with a different number of realizations could also cause changes in the CCS values, particularly if the number of realizations is much lower/higher than 500. This is because a $P_0$ value fails to meet our CCS condition if even a single realization (at that $P_0$-value) experiences a fade-out. CCS values are initial population sizes ($P_0$-values), the actual population size at 1 or 5 years will differ due to births, deaths, and the stochastic nature of our model. To calculate the proportion of the population infected (after 1 and 5 years) we divide the total number of individuals who were ever infected (current infections, individuals who died due to COVID-19, individuals who recovered from COVID-19, and individuals who recovered from COVID-19 but subsequently died for unrelated reasons) by the total number of individuals who have ever existed in that population (both currently living and dead) by that time-step ($t = 365$ days, $t = 1825$ days respectively) and thus could potentially have become infected.

**Table S1. Model parameters.** Values drawn from empirical data where possible.

| Parameter | Description | Baseline value | Source |
|---|---|---|---|
| $b$ | Probability of giving birth | $5.1 \times 10^{-5}$/day | [6] |
| $d$ | Probability of death (non-COVID-19 related) | $2.1 \times 10^{-5}$/day | [6] |
| $m$ | Probability of a symptomatic individual dying due to COVID-19 | 0.0066 | [1] |
| $\tau_I$ | Symptomatic testing probability | 0.3/day | [7] |
| $\alpha$ | Probability of $E \to A$ transition | 0.4/day | [8, 9] |
| $\sigma$ | Probability of $A \to I$ transition | 0.4/day | [8, 9] |
| $\rho$ | Probability of $I \to R$ and $A \to R$ transitions | 0.67/day | [8, 9] |
| $s$ | Super-spreader parameter | 0.2 | [2] |
| $\pi$ | Proportion of individuals who are asymptomatic | 0.2 | [10] |
| $w$ | Proportion of contacts in schools and workplaces | 0.45 | [11] |
| $\omega$ | Risk perception proportionality constant | $4.4 \times 10^4$ | [7] |
| $\varepsilon$ | Efficacy of NPIs | 0.85 | |
| $\beta_0^P$ | Transmission probability, pre-symptomatic | 0.43/day | $\beta_0^P = 0.5\beta_0^I$, [3, 4] |
| $\beta_0^A$ | Transmission probability, asymptomatic | 0.43/day | $\beta_0^A = 0.5\beta_0^I$, [3, 4] |
| $\beta_0^I$ | Transmission probability, symptomatic | 0.86/day | [3, 4] |
| $\xi$ | Proportion of mass-action incidence | 0 | [12, 13] |
| $\eta$ | Adherence to self-isolation | 0.8 | [14, 15] |
| $\gamma$ | Threshold prevalence which triggers a closure | $1.5 \times 10^{-4}$ | [7] |
| $\delta_C$ | Length of a closure period (i.e. minimum closure duration) | 30 days | [7] |

### Supplementary figures

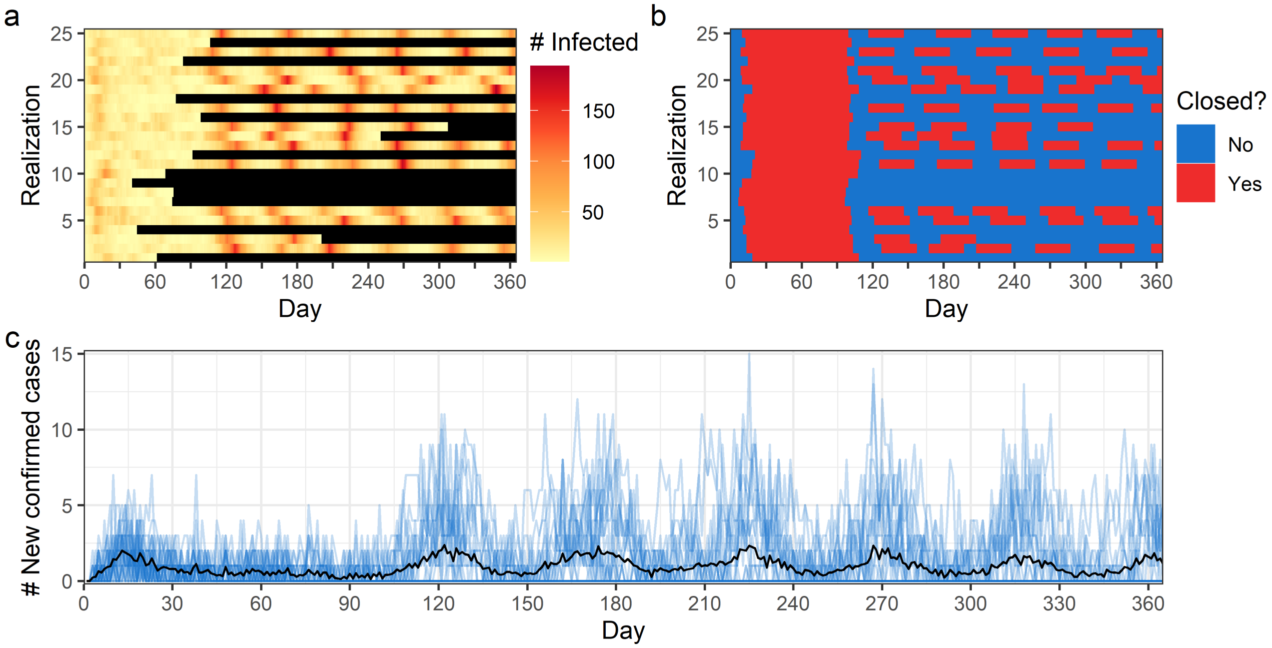


**Figure S1. Effective strategy where many populations quickly eradicate the pathogen due to strong risk aversion within the populace.** Figure panels show (a) number of infected individuals within the population (black indicates zero infected) (b) whether closures are in place and daily number of (c) new confirmed cases. In (a) and (b) each row corresponds to a single realization, while in (c) blue lines indicate each realization, with the black line indicating the mean outcome across all realizations. Model parameter settings are shown in Table S1, except for $\omega = 4.4 \times 10^5$.

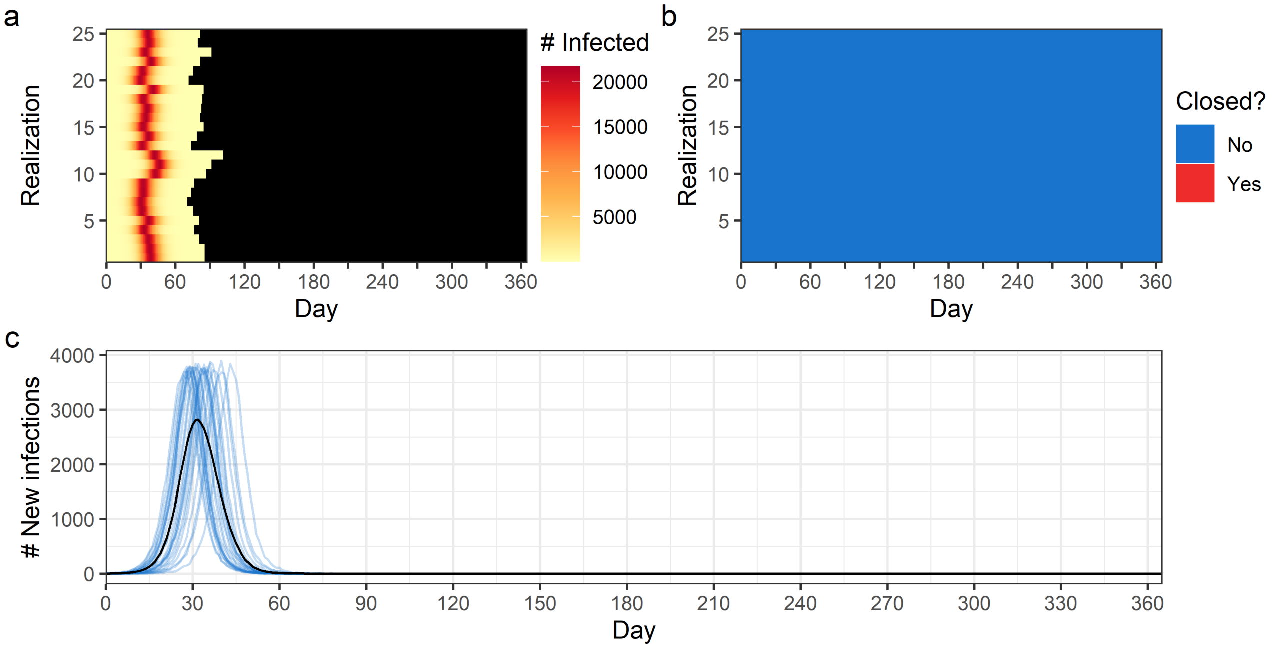


**Figure S2. Rapid burn out occurs when non-pharmaceutical interventions are not introduced.** Figure panels show (a) number of infected individuals within the population (black indicates zero infected) (b) whether closures are in place and daily number of (c) new infections. In (a) and (b) each row corresponds to a single realization, while in (c) blue lines indicate each realization, with the black line indicating the mean outcome across all realizations. Model parameter settings are shown in Table S1, except for $\tau_I = 0$. This setting prevents any individual or collective NPIs from being employed by the simulated populations, as NPIs are implemented based on known case prevalence.

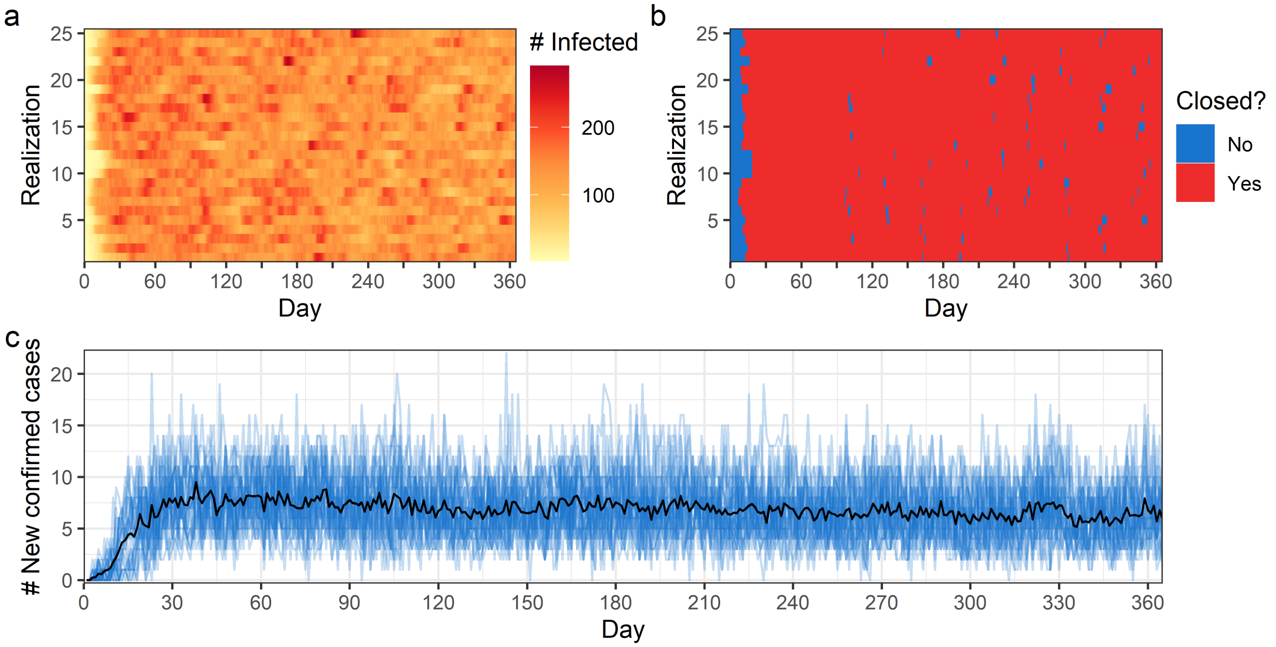


**Figure S3. Ineffective strategy where new cases decrease extremely slowly due to low risk aversion within the populace.** Figure panels show (a) number of infected individuals within the population (black indicates zero infected) (b) whether closures are in place and daily number of (c) new confirmed cases. In (a) and (b) each row corresponds to a single realization, while in (c) blue lines indicate each realization, with the black line indicating the mean outcome across all realizations. Model parameter settings are shown in Table S1, except for $\omega = 4.4 \times 10^3$.

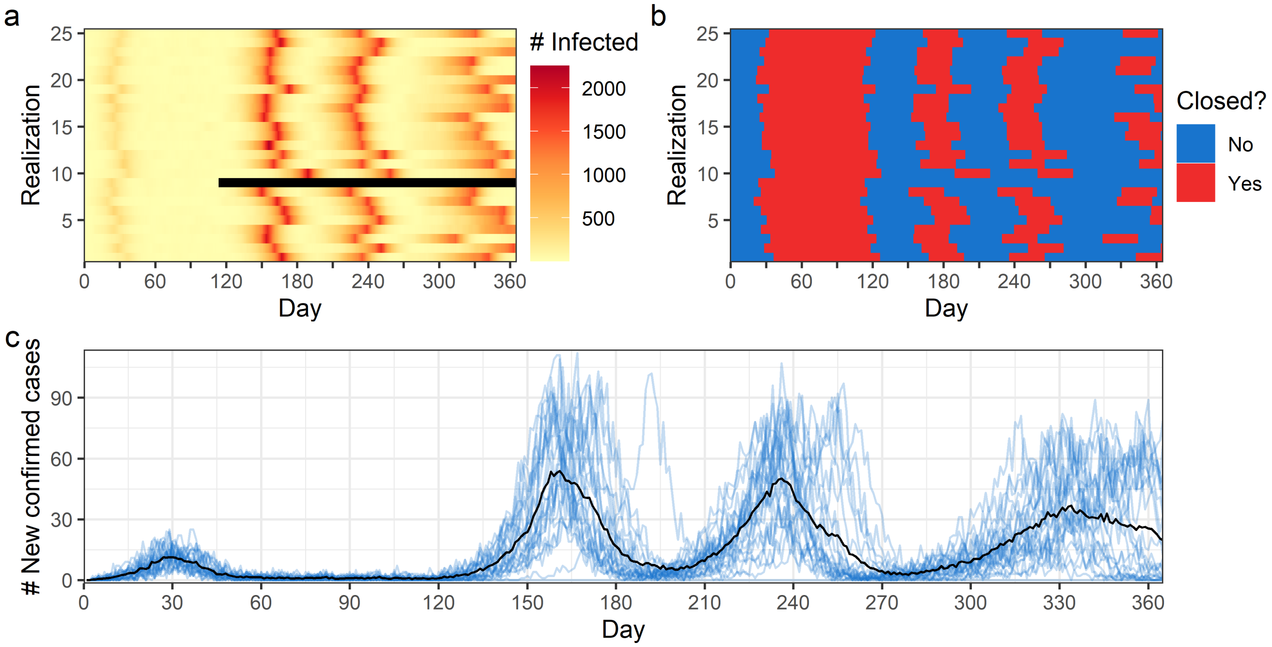


**Figure S4. Ineffective strategy where many individuals become infected because closures are not triggered until case prevalence is high.** Figure panels show (a) number of infected individuals within the population (black indicates zero infected) (b) whether closures are in place and daily number of (c) new confirmed cases. In (a) and (b) each row corresponds to a single realization, while in (c) blue lines indicate each realization, with the black line indicating the mean outcome across all realizations. Model parameter settings are shown in Table S1, except for $\gamma = 2 \times 10^{-3}$.

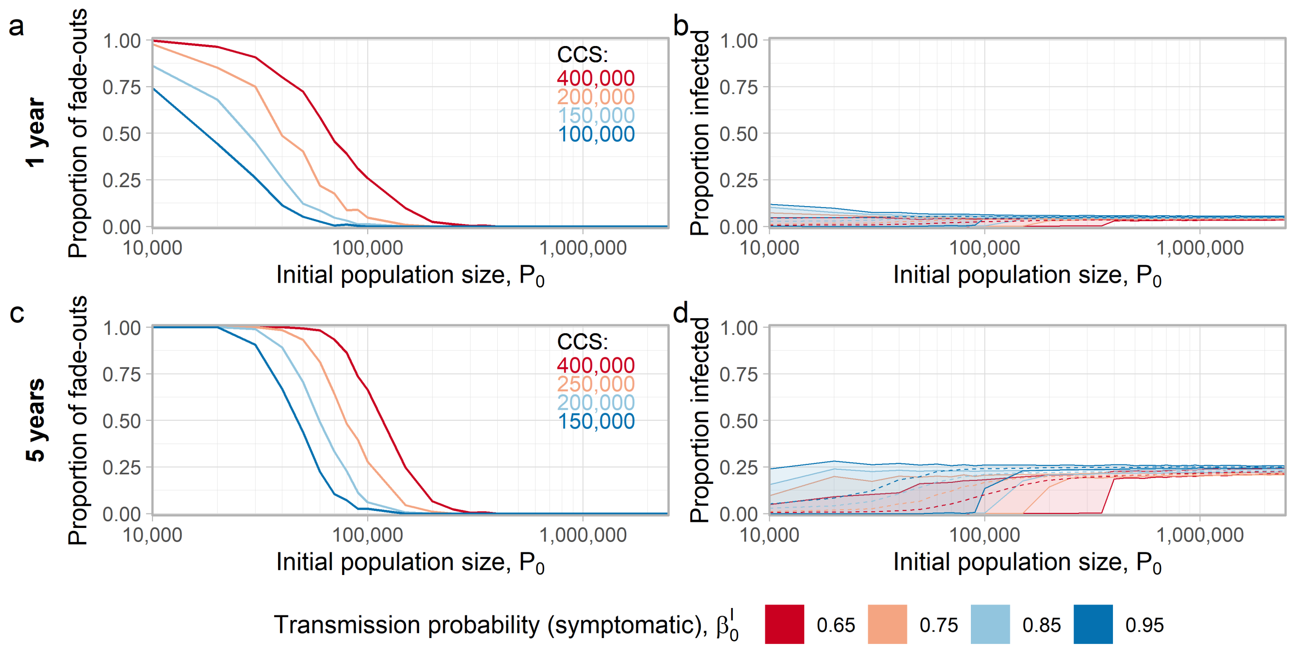


**Figure S5. Higher transmission probability (symptomatic) reduces the critical community size, increases the proportion of individuals who become infected.** Figure panels show (a) the proportion of fade-outs and (b) the mean proportion of the population infected (dashed lines) after 1 year and 5 years in (c) and (d). In (a) and (c) inset panels indicate critical community sizes, and in (b) and (d) ribbons display minimal and maximal values across all realizations. Settings for all parameters (except $\beta_0^I$) are shown in Table S1.

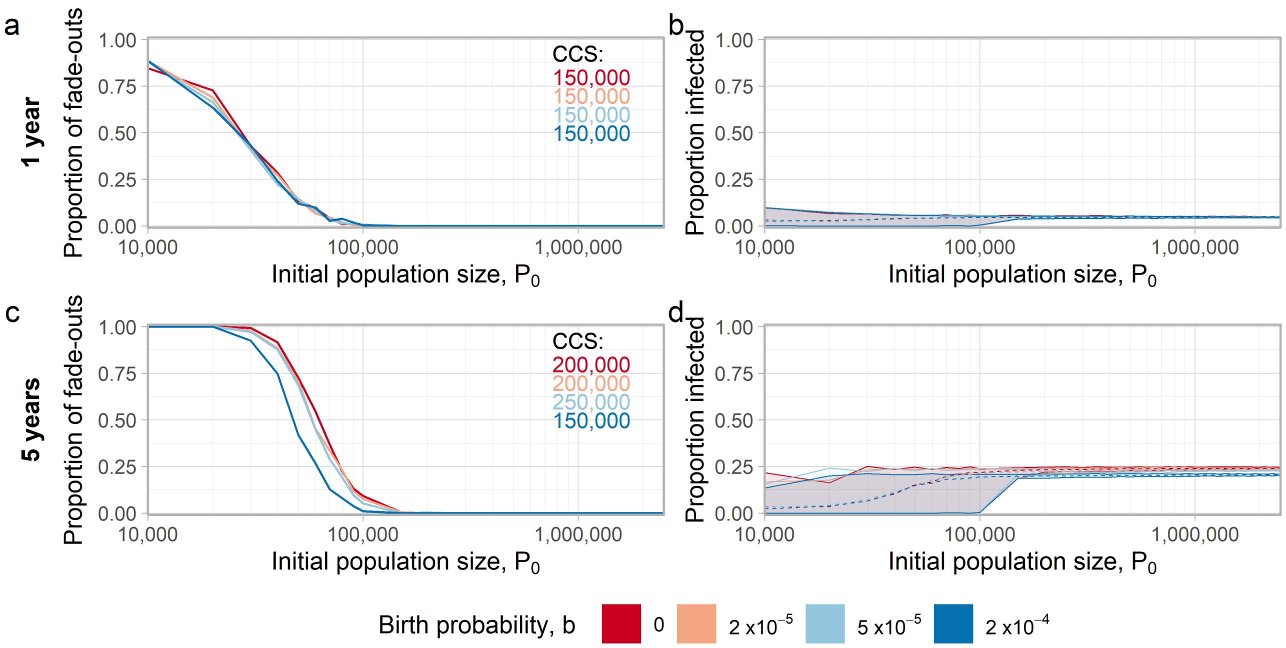


**Figure S6. Birth rates have little effect on the critical community size and proportion of the population who become infected.** Figure panels show (a) the proportion of fade-outs and (b) the mean proportion of the population infected (dashed lines) after 1 year. Results after 5 years are shown in (c) and (d). In (a) and (c) inset panels indicate critical community sizes, and in (b) and (d) ribbons display minimal and maximal values across all realizations. Settings for all parameters (except $b$) are shown in Table S1.

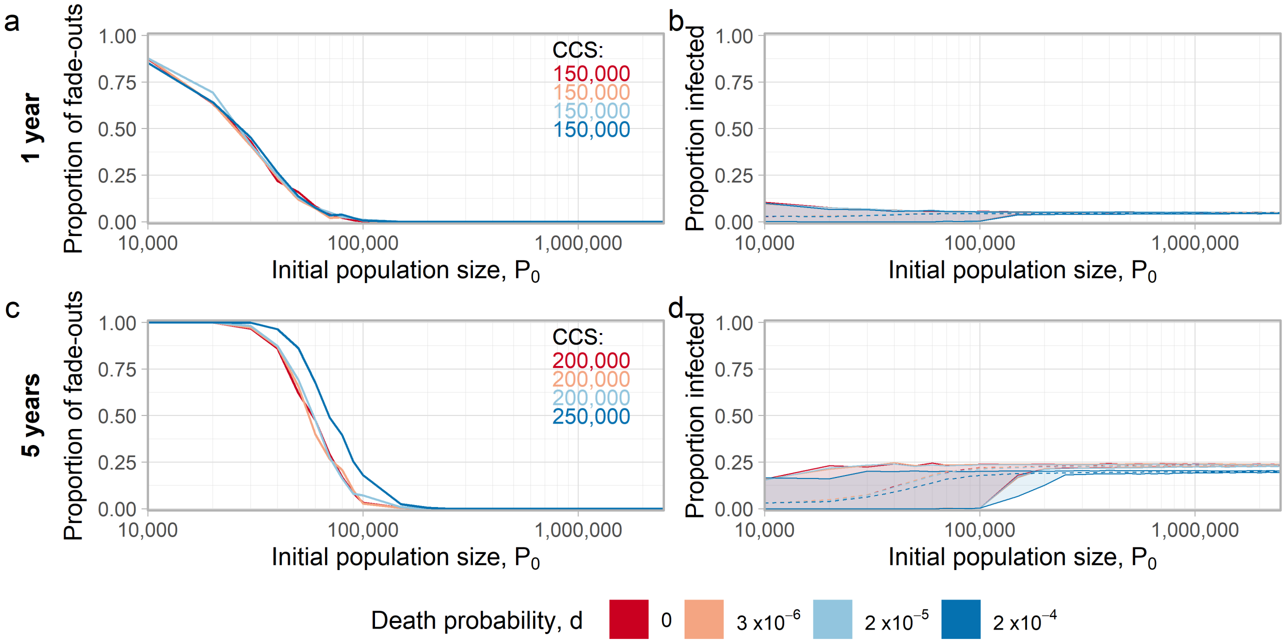


**Figure S7. Death rates have little effect on the critical community size, but higher death rates slightly lower the proportion of the population who become infected.** Figure panels show (a) the proportion of fade-outs and (b) the mean proportion of the population infected (dashed lines) after 1 year. Results after 5 years are shown in (c) and (d). In (a) and (c) inset panels indicate critical community sizes, and in (b) and (d) ribbons display minimal and maximal values across all realizations. Settings for all parameters (except $d$) are shown in Table S1.

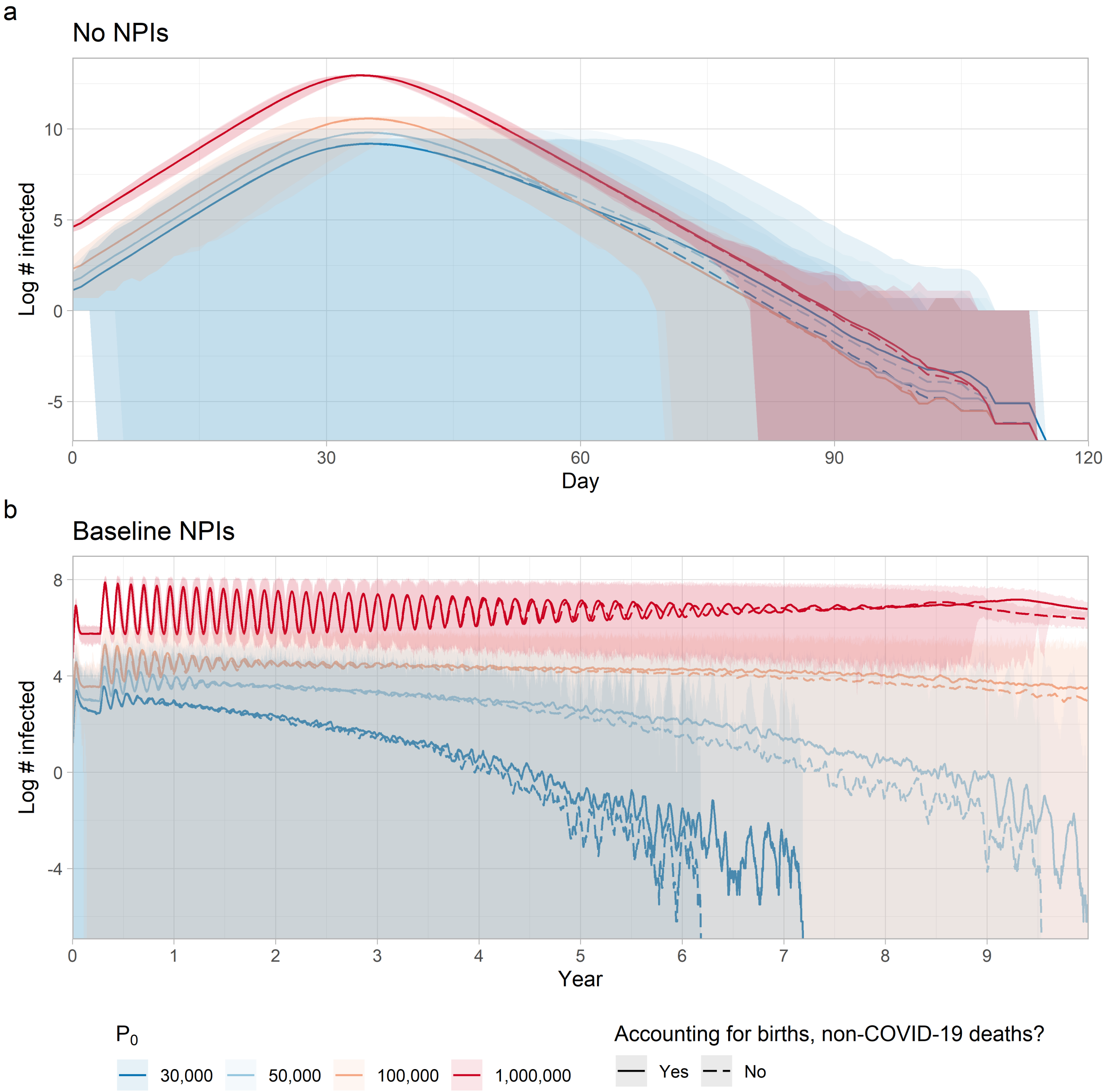


**Figure S8. In the presence of NPIs, accounting for births and deaths results in longer times to fade-out.** Figure panels show (a) the proportion of fade-outs and (b) the mean proportion of the population infected (dashed lines) after 1 year. Results after 5 years are shown in (c) and (d). In (a) and (c) inset panels indicate critical community sizes, and in (b) and (d) ribbons display minimal and maximal values across all realizations. Settings for all parameters are shown in Table S1, with $b = d = 0$ in the scenario where births and non-COVID-19 deaths are not accounted for.

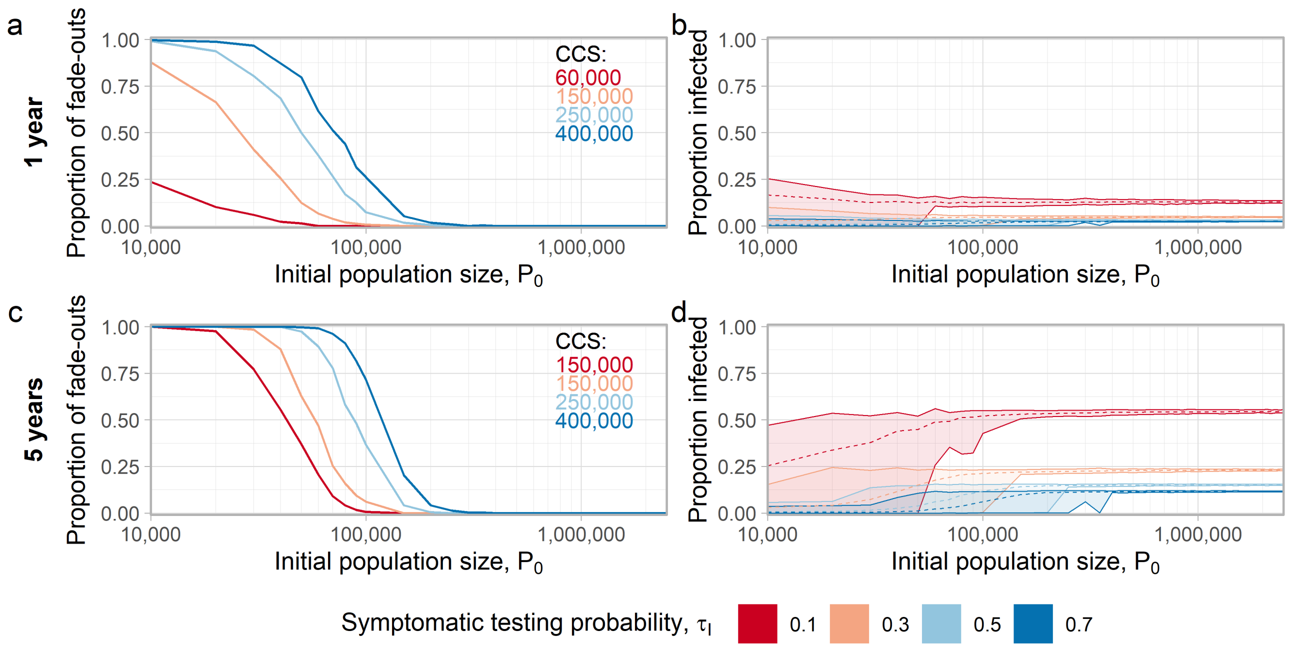


**Figure S9. Higher testing rate increases the critical community size, reduces the proportion of individuals who become infected.** Figure panels show (a) the proportion of fade-outs and (b) the mean proportion of the population infected (dashed lines) after 1 year. Results after 5 years are shown in (c) and (d). In (a) and (c) inset panels indicate critical community sizes, and in (b) and (d) ribbons display minimal and maximal values across all realizations. Settings for all parameters (except $\tau_I$) are shown in Table S1.

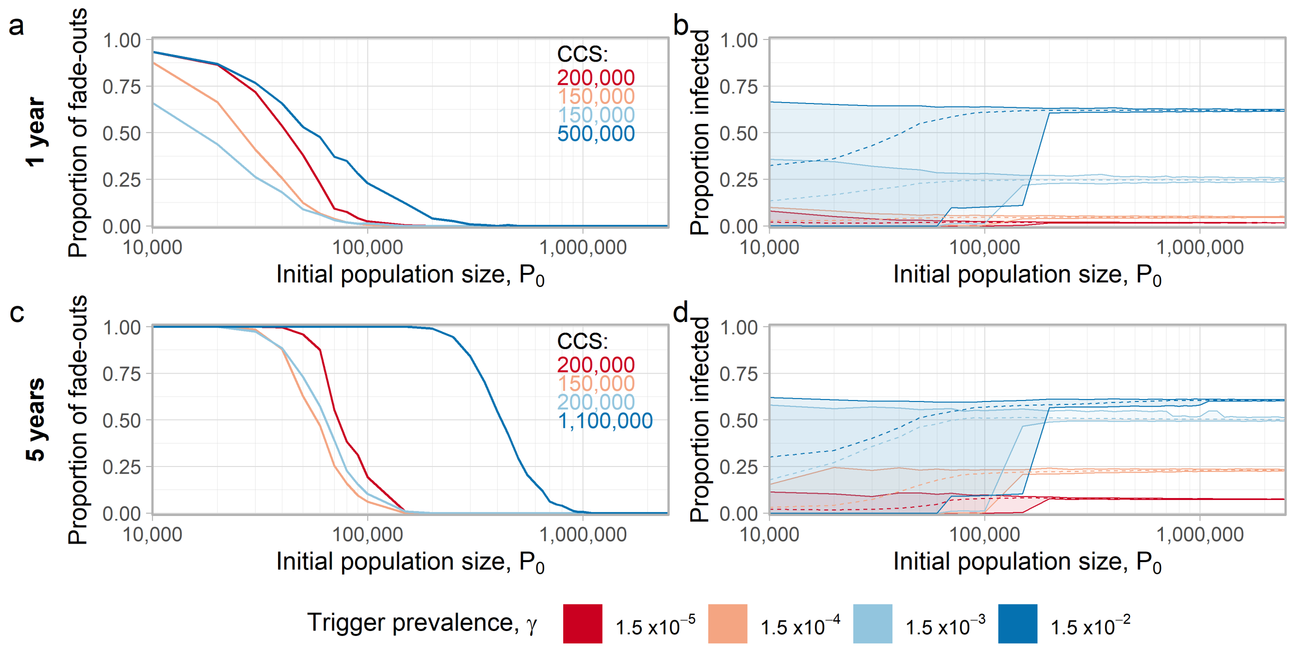


**Figure S10. Lower trigger prevalence reduces the proportion of the population that becomes infected, may increase CCS.** Figure panels show (a) the proportion of fade-outs and (b) the mean proportion of the population infected (dashed lines) after 1 year. Results after 5 years are shown in (c) and (d). In (a) and (c) inset panels indicate critical community sizes, and in (b) and (d) ribbons display minimal and maximal values across all realizations. Settings for all parameters (except $\gamma$) are shown in Table S1.

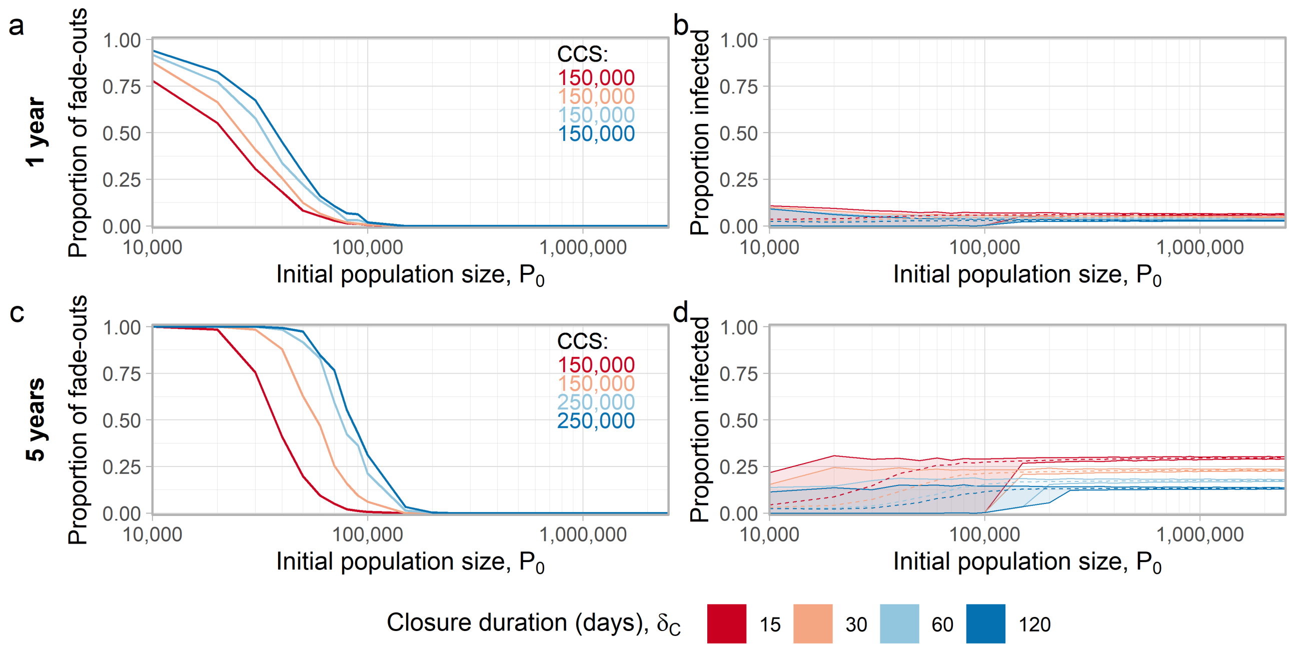


**Figure S11. Committing to longer closures reduces the proportion of individuals infected, increases critical community size.** Figure panels show (a) the proportion of fade-outs and (b) the mean proportion of the population infected (dashed lines) after 1 year. Results after 5 years are shown in (c) and (d). In (a) and (c) inset panels indicate critical community sizes, and in (b) and (d) ribbons display minimal and maximal values across all realizations. Settings for all parameters (except $\delta_C$) are shown in Table S1.

### References


1. Robert Verity, Lucy C Okell, Ilaria Dorigatti, Peter Winskill, Charles Whittaker, Natsuko Imai, Gina Cuomo-Dannenburg, Hayley Thompson, Patrick GT Walker, Han Fu, et al. Estimates of the severity of coronavirus disease 2019: a model-based analysis. *The Lancet infectious diseases*, 2020.

2. James O Lloyd-Smith, Sebastian J Schreiber, P Ekkehard Kopp, and Wayne M Getz. Superspreading and the effect of individual variation on disease emergence. *Nature*, 438(7066):355–359, 2005.

3. Xi He, Eric HY Lau, Peng Wu, Xilong Deng, Jian Wang, Xinxin Hao, Yiu Chung Lau, Jessica Y Wong, Yujuan Guan, Xinghua Tan, et al. Temporal dynamics in viral shedding and transmissibility of covid-19. *Nature medicine*, 26(5):672–675, 2020.

4. Joe Hilton and Matt J. Keeling. Estimation of country-level basic reproductive ratios for novel coronavirus (covid-19) using synthetic contact matrices. *medRxiv*, 2020.

5. Herbert W Hethcote. The mathematics of infectious diseases. *SIAM review*, 42(4):599–653, 2000.

6. UN DESA, Population Division. World population prospects 2019, online edition, 2019. Data retrieved on June 9, 2020 from `https://population.un.org/wpp/Download/Standard/Population/`.

7. Vadim A Karatayev, Madhur Anand, and Chris T Bauch. Local lockdowns outperform global lockdown on the far side of the covid-19 epidemic curve. *Proceedings of the National Academy of Sciences*, 117(39):24575–24580, 2020.

8. Hiroshi Nishiura, Natalie M Linton, and Andrei R Akhmetzhanov. Serial interval of novel coronavirus (covid-19) infections. *International journal of infectious diseases*, 2020.

9. Lauren Tindale, Michelle Coombe, Jessica E Stockdale, Emma Garlock, Wing Yin Venus Lau, Manu Saraswat, Yen-Hsiang Brian Lee, Louxin Zhang, Dongxuan Chen, Jacco Wallinga, et al. Transmission interval estimates suggest pre-symptomatic spread of covid-19. *MedRxiv*, 2020.

10. Kenji Mizumoto, Katsushi Kagaya, Alexander Zarebski, and Gerardo Chowell. Estimating the asymptomatic proportion of coronavirus disease 2019 (covid-19) cases on board the diamond princess cruise ship, yokohama, japan, 2020. *Eurosurveillance*, 25(10):2000180, 2020.

11. Bureau of Labor Statistics. American time use survey - 2018 results, 2018. Retrieved on June 16, 2020 from `https://www.bls.gov/news.release/pdf/atus.pdf`.

12. MCD Jong, O Diekmann, and H Heesterbeek. How does transmission of infection depend on population size? epidemic models. *Publication of the Newton Institute*, pages 84–94, 1995.

13. Janis Antonovics, Yoh Iwasa, and Michael P Hassell. A generalized model of parasitoid, venereal, and vector-based transmission processes. *The American Naturalist*, 145(5):661–675, 1995.

14. FA Soud, MM Cortese, AT Curns, PJ Edelson, RH Bitsko, HT Jordan, AS Huang, JM Villalon-Gomez, and GH Dayan. Isolation compliance among university students during a mumps outbreak, kansas 2006. *Epidemiology & Infection*, 137(1):30–37, 2009.

15. Joel Hellewell, Sam Abbott, Amy Gimma, Nikos I Bosse, Christopher I Jarvis, Timothy W Russell, James D Munday, Adam J Kucharski, W John Edmunds, Fiona Sun, et al. Feasibility of controlling covid-19 outbreaks by isolation of cases and contacts. *The Lancet Global Health*, 2020.
